# Tracking COVID-19 Cases and Deaths in the United States: Distribution of Events by Day of Pandemic

**DOI:** 10.1101/2021.08.30.21262851

**Authors:** Andrew Moore, Mingdong Lyu, Randolph Hall

## Abstract

In this paper, we analyze the progression of COVID-19 in the United States over a nearly one-year period beginning March 1, 2020, with a novel metric representing the partial-average day-of-event, where events are new cases and new deaths. The metric is calculated as a function of date and location to illustrate patterns of disease, showing growing or waning cases and deaths. The metrics enable the direct comparison of the time distribution of cases and deaths, revealing data coherence and how patterns varied over a one-year period. We also compare different methods of estimating actual infections and deaths to better understand on the timing and dynamics of the pandemic by state. We used three example states to graphically compare metrics as functions of date and also compared statistics derived from all 50 states. Over the period studied, average case day and average death day vary by two to five months among the 50 states, depending on data source, with the earliest averages in New York and surrounding states, as well as Louisiana. The average day of death has preceded the average day of case in Centers for Disease Control (CDC) data for most states and most dates since June of 2020. In contrast, “COVID-19 Projections” more closely align deaths and cases, which are similarly distributed.

## Introduction

In this paper, we investigate differences in COVID-19’s spread in the United States by examining the time distribution of events, where an event is either a new case or new death attributed to COVID-19. We focus on the cumulative distribution of events within time-periods along with the average time of events occurring within time-periods. We compare three data sources, including the Centers for Disease Control’s data on reported events and two other sources that have used inference methods to estimate actual infections and deaths.

Our examination provides normalized metrics as to when the disease accelerated and decelerated by location and how the disease patterns varied among the 50 United States. As the novel Coronavirus disease-19 (COVID-19) pandemic progressed across the United States in 2020 and early 2021, the rates of cases and deaths varied by week and month among states. While at the national level the United States faced three major case waves (Spring 2020, Summer 2020, and Winter 2020/21) during our period of study, the time distribution of cases and deaths has varied by location. Very early in the pandemic, large urban areas, especially in the northeast, had the highest rates of cases and deaths. New York City was the pandemic’s epicenter, registering 203,000 laboratory-confirmed cases in the first three months of the pandemic (1). However, by July 2020, southern states, such as Florida, Texas, and Arizona, became hot spots for the pandemic. By late summer and early fall, Midwestern states, such as North Dakota, South Dakota, and Iowa, surged in cases and deaths (2). Rural areas were hardest hit during this period. November through January represented a national surge in cases, when many states recorded new highs in cases. States with lower case rates in the summer, like New York and New Jersey, had surges again. In December of 2020, 21 states registered at least 2,000 new cases per 100,000 residents, and twenty-six states had rates between 1,000 and 2,000 new cases per 100,000 residents (3). By February, case rates started dropping off across the United States.

Our metrics provide a novel assessment of the progression of the disease, offering a way to examine the current state of the disease in the context of the history of cases and deaths. In the following section, we review prior research examining patterns of COVID-19 in the United States. We next introduce our methodology for developing metrics for tracking disease-related events and illustrate these metrics for an example of geometric growth and decay. Then we present results, including graphs representing example states and a comparison of statistics for all 50 states. We end with conclusions and discussion.

## Literature Review

As the novel coronavirus spread worldwide, cases, hospitalizations and deaths have been studied with epidemiological models, characterizing the COVID-19 epidemic, forecasting transmission, and evaluating pharmaceutical and non-pharmaceutical intervention. Mathematical methods, such as compartmental models, statistical models, general machine learning models, and agent-based models have been applied to simulate, characterize, and forecast COVID-19 (4; 5; 6; 7; 8).

Early mathematical models of COVID-19 updated previous disease models to consider the unique characteristic of COVID-19 (9), including updating older SIR models to consider hospitalized and undetected infections (9; 10; 11). Other research focused on risk factors and epidemiological characteristics of COVID-19 patients, such as fevers and coughing (12; 13; 14). Early research focused on Wuhan, prior to when disease spread to other countries. Later research focused on how the outbreak dynamics had differed in countries such as China and the United States (15). This type of research started analyzing localities because of differing transmission rates, which may be related to human behavior, living conditions and environmental factors. Furthermore, epidemiological research started looking at transmission rates in different settings (i.e., homes and work) in other countries.

To inform intervention efforts, state and local health departments have used various indicators to identify the changes in the number of cases, hospitalization, and deaths. For example, the Kentucky Department for Public Health (KDPH) adopted a composite syndromic surveillance data, number of new cases, number of COVID-19 associated deaths, health care capacity data, and contact tracing capacity as five indicators to assess the state-level COVID-19 status (16). From empirical data, they scored each of the five indicators using a 3-point scale (3= excellent, 2 =moderate, 1 = poor), and then combined the five scores with equal weights to a composite state-level COVID-19 status by a 5-point rating system. KDPH found that during May 19 – July 15, 2020, the Kentucky composite COVID-19 status worsened. Similarly, King County in Washington state published a Key Indicators dashboard to provide an overview of how they are doing in important areas: disease activity, testing and healthcare system status (17). King County calculated the 7-day average cases per capita, 7-day average hospitalization per capita, 7-day average deaths per capita, effective reproduction number, and percentage of occupied hospital beds. The performance in each area could be assessed by the comparison of the five indicators with the settled target. These indicators provided a plain language assessment to facilitate reopening decisions.

In addition, health care indicators can help compare the healthcare outcomes across populations. For example, cumulative death counts are associated with demographic characteristics. Heuveline and Tzen proposed three measures – the crude death rate, the age-standardized death rate, and life expectancy at birth – to compare status over time (18). They disaggregate the population into smaller administrative units with respect to age and sex to provide more meaningful comparisons.

During the pandemic, it has been apparent that data and predictions influence policy decisions aimed at lessening the impact of COVID-19, yet suffer from significant uncertainty. In this environment, it is difficult to develop the most effective interventions, and instill public confidence that the most effective interventions have been selected (19; 20).

Our research focuses on a novel way to track data on cases and deaths. We focus on the time distribution of cases and deaths as a standardized indicator for the time-varying dynamic of the pandemic, showing when it is accelerating or decelerating in the United States on a state level. We calculate and compare partial averages by day to quantify the progression of the disease. We focus on state-level data to model how the progression of the disease has affected each state of the country at different times. Finally, we assess three data sources to understand how different estimation methods (relative to official counts of reported cases and deaths) affect how the pandemic appears to have developed in each state.

## Methodology

Our research analyzes the time distribution of reported (and estimated) cases and deaths attributed to COVID-19 in the United States, beginning on March 1 of 2020. We focus on the time distribution of reported events, as indicators of the pandemic’s progression by location. In this section we develop a methodology for analyzing event statistics, focusing on partial averages, as a function of date, along with the cumulative distribution of events in the study period. An idealized geometric progression is used to illustrate properties of time partial-averages. Understanding the properties of the idealized model will help to understand empirical results, which is the focus of Section 4.

### Computing Partial Averages by Day

We consider a process where events are tracked over time by counting the number occurring on each day. Our focus is on cases (and estimated infections) and deaths, but the concepts generalize to any process where events are counted by time increment. We seek to track partial averages, representing the average day of events occurring on day t or earlier. Let:

*A*(*t*) = average event day, for events occurring on day *t* or earlier.

Day 1 (*t* = 1) represents the day of the first recorded event. By definition, *A*(1) must equal 1. *A*(*t*) is a non-decreasing function, with these properties:

*A*(*t* + 1) *> A*(*t*) if new events occur on day *t* + 1

*A*(*t* + 1) = *A*(*t*) if no new events occur on day *t* + 1

Let:

*T* = day of last recorded event

*P* (*t*) = proportion of events that occurred on day *t* or earlier

By definition, *P* (*T*) = 1, as all events must occur before day *T* . Let:

*p*(*i*) = proportion of events that occur on day i

Then:

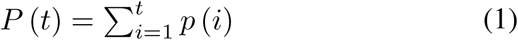

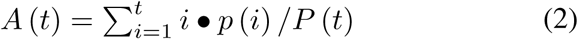

*A*(*t* + 1) can be computed as a weighted average, as follows:

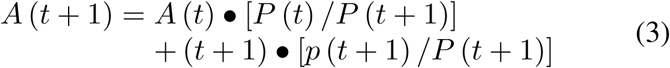

Let Δ(*t* + 1) represent the change in average event time from day t to day t+1:

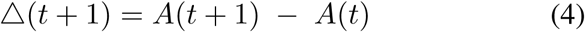

Δ (*t* + 1) can be derived algebraically from Eq.(3), resulting in:

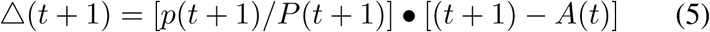

Eq. 5 offers insight into how rapidly *A*(*t*) changes as a consequence of *p*(*t* + 1), rising fastest in time periods when the probability of an event exceeds the rate of events in the preceding days, such as when events are occurring at an accelerating rate. Another insight is that *A*(*t*) can grow particularly fast in a second or later wave of a pandemic. After an initial wave, *A*(*t*) may remain stable for a period of time, as *p*(*t* + 1) is small. If a pandemic re-emerges, then *A*(*t*) may grow rapidly, both because *p*(*t* + 1) is large and because *t* + 1 is now much greater than *A*(*t*) (owing to the prior lull when *A*(*t*) was increasingly slowly).

### Geometric Growth and Decay Example

To understand patterns in empirical data from COVID-19, we first explore the characteristics of the idealized scenarios of geometric growth and decay, characterized by the function:

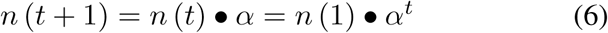

where

*n* (*t*) = number of events occurring in day t

*α* = rate of growth (or decay) in events.

Geometric growth occurs when *α* is greater than 1, geometric decay when *α* is less than 1, and events occur at a constant rate when *α* equals 1. Let:

*N* (*t*) = cumulative events from day 1 to day *t*

Then:

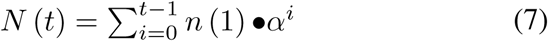

As a geometric progression, N(t) can be expressed:

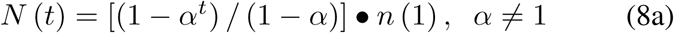

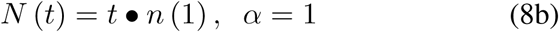

As defined in Eq. 4, Δ(*t* + 1) is the change in the average event day from day *t* to day *t* + 1. Substituting *n*(*t* + 1)*/N* (*t* + 1) from above for the ratio *p*(*t* + 1)*/P* (*t* + 1):

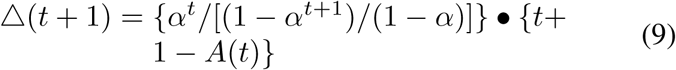

In the special case where *α >* 1 (geometric growth), Δ(*t* + 1) is an increasing function, approaching the limit:

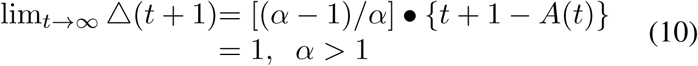

And further,

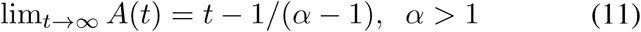

In the special case where *α <* 1 (geometric decay), Δ(*t* + 1) is a decreasing function, approaching the limit:

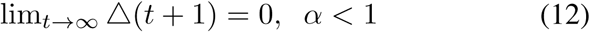

and, further,

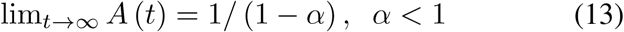

Last, in the special case where *α* = 1 (constant), Δ(*t* + 1) = .5 for all values of t, and thus *A*(*t*) is simply (1 + *t*)*/*2.

The properties of *A*(*t*) are illustrated in Figure 1 for example rates of growth (10%, *α* = 1.1; 5%, *α*=1.05) and decay (−5%, *α* =.95), as well as constant (0%, *α* = 1.0). For instance, for 10% growth, *A*(*t*) approaches t-10 (equalling *t* − 1*/*(*α* − 1)), and for 5% decay, *A*(*t*) approaches 20, equalling 1*/*(1 − *α*)).

**Figure 1.**
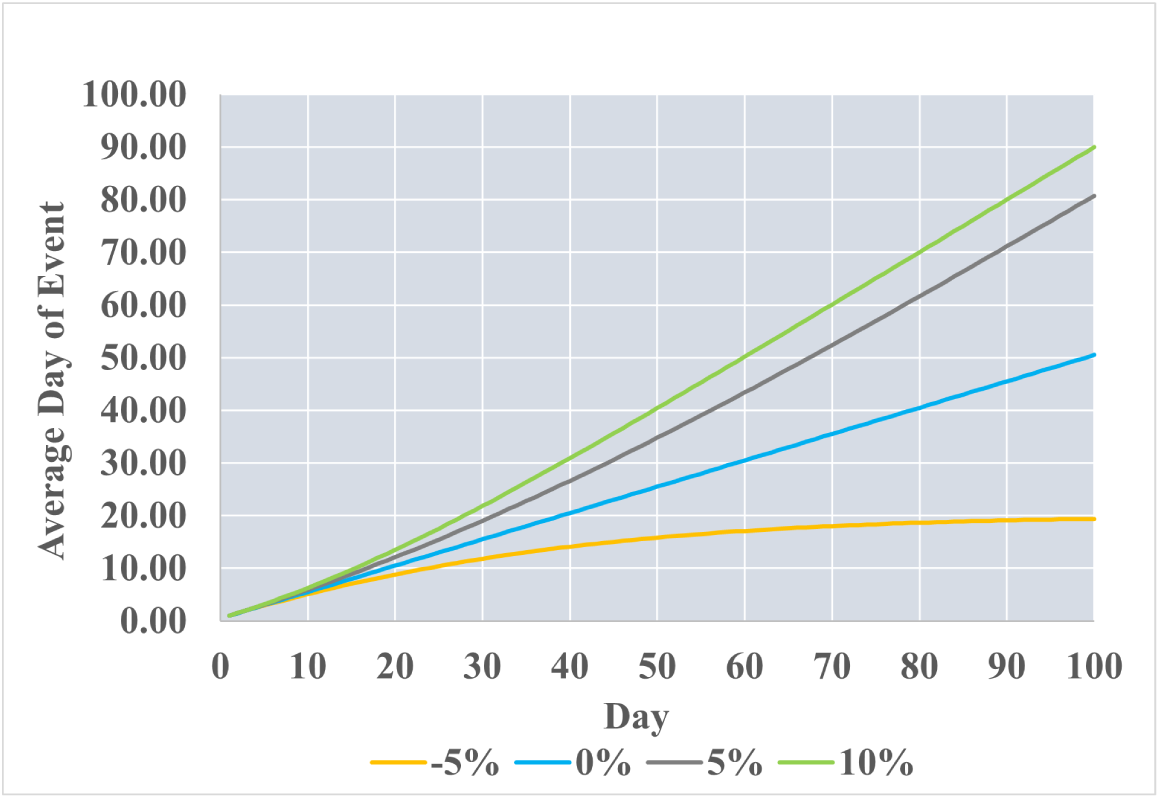
Average Day of Event as Function of Day (geometric growth or decay, by % daily change)

For another metric, we introduce:

H(t) = days elapsed since average event on day *t* = *t* − *A*(*t*).

Figure 2 illustrates the limiting properties of H(t), which approaches the constant 1*/*(*α* − 1) under geometric growth; approaches *t* − 1*/*(1 − *α*) under geometric decay; and equals *t/*2 − 1*/*2 when *α* = 1. Last, Figure 3 plots *H*(*t* + 1) − *H*(*t*), illustrating again that for geometric growth, H(t) approaches a limiting value as t becomes large, yet approaches a constant growth rate of 1 for geometric decay.

**Figure 2.**
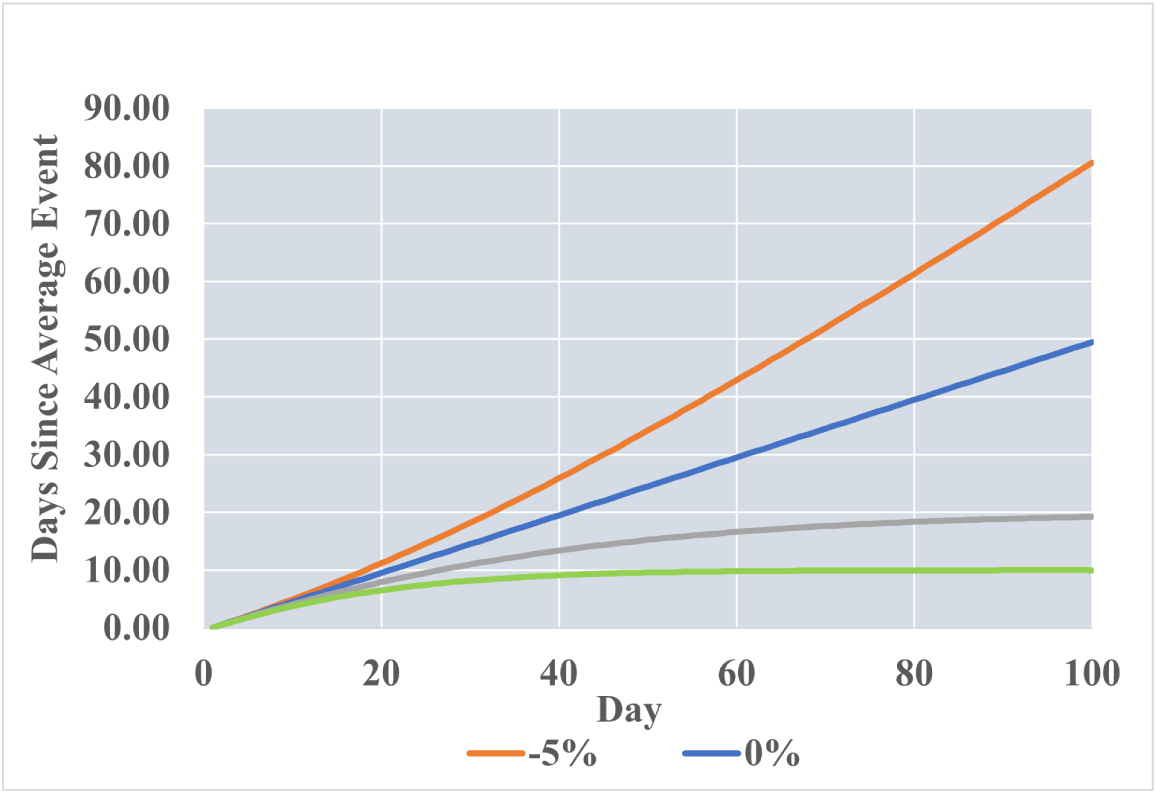
Days Since Event as Function of Day (geometric growth or decay, by % daily change)

**Figure 3.**
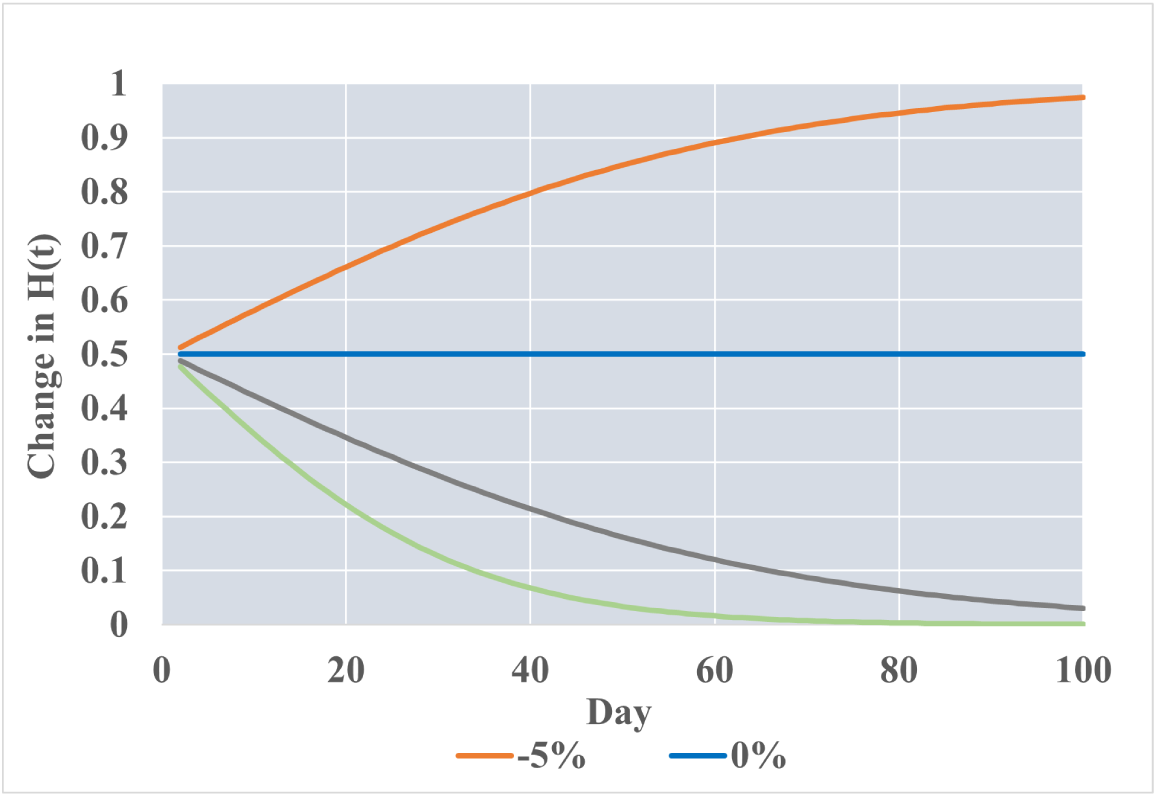
Daily Change in Average Days Since Event as Function of Day (geometric growth or decay, by % daily change)

As we examine empirical data in individual states in Section 4, we will look for signals of geometric growth (when the pandemic is accelerating) or decay (when the pandemic is waning) through our examination of *A*(*t*) for cases and deaths. As mentioned, Δ(*t* + 1) never exceeds one for the idealized geometric model. As will be seen, Δ(*t* + 1) can exceed one when events occur in multiple waves, as has been the case for COVID-19. We will refer to this as super-geometric growth.

### Source Data and Indicators

We compared data from three sources: CDC Reported Data, Covidestim estimated data, and COVID-19 Projections estimated data. Estimated data attempt to correct for errors in reporting, which might, for example, include infections that were never detected through testing.

CDC reported data consists of two metrics for recorded cases and deaths: confirmed case or death and probable case or death (4). The CDC defines a confirmed case or death as met by confirmed laboratory evidence for COVID-19. Furthermore, the CDC defines a probable case or death as meeting one of three standards (12):

- Clinical criteria and epidemiological evidence with no confirmed laboratory testing
- Laboratory evidence and either clinical criteria or epidemiological evidence
- Vital records with no confirmatory laboratory testing We used the total combined count of confirmed and probable for cases and deaths. The CDC further explains how the reported data can fluctuate, due to:
- Jurisdictions reclassifying probable cases,
- Counts being revised as records are finalized,
- Jurisdictions having different reporting time intervals and methodologies for reporting cases and deaths,
- Delays in reporting and testing.

We compared metrics for CDC data to two estimated data sources: Covidestim and COVID-19 Projections. Covidestim is an experimental methodology using Bayesian evidence synthesis to adjust reported data: accounting for asymptomatic infections, undercounting due to lack of availability of testing, and delays in case and death counts (21). The model was run every 28 days, given the lag time for observed data, and is parameterized by four health states: asymptomatic, symptomatic, severe, and death (21).

COVID-19 Projections is an experimental “nowcasting” model that aims to standardize the test positivity and estimate the true incidence of COVID-19. The model adjusts the test positivity by taking the average ratio for each date and applying this ratio to states that report “unique people” (22). Next, the model estimates the prevalence ratio by using the positivity rate and date to estimate true infections (22). Finally, the model estimates the number of infections by parametrizing the prevalence ratio, positivity rate, and confirmed cases using a 7-day moving average (22). For our purposes, we will use COVID-19 Projections estimated infections and total deaths.

Our analysis is based on the period March 1, 2020 through February 21, 2021, with the end date matching the end date of COVID-19 Projections’ estimation of infections. Data were downloaded on these dates: Covidestim, June 25, 2021; COVID-19 Projections, June 26, 2021; CDC, June 25, 2021.

## Results

We examined case and death statistics/estimates for COVID-19 for all 50 of the United States for the three data sources CDC, Covidestim and COVID-19 Projections. For each state and data source, we calculated *A*(*t*) and *P* (*t*) for deaths and cases. We also compared statistics as follows. Let:

*A*_*d*_(*t*) = average death day for events occurring on day t or earlier

*A*_*c*_(*t*) = average case day for events occurring on day t or earlier.

We then computed the difference between average death day and average case day:

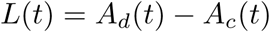

Though for any individual, deaths must follow initial infection, we will see that *L*(*t*) is not necessarily positive.

We also calculated the ratios:

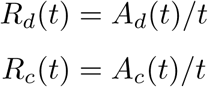

These ratios are naturally close to one at the onset of a pandemic (i.e., average day of event is similar to the current day). They remain close to one if cases and/or deaths grow geometrically and they drop toward zero if cases and/or deaths decay geometrically. In this manner, the ratios are indicators of the progression of a pandemic.

### Disease Patterns in Example States

For illustration, graphs are presented here for three states – California, New York, Minnesota – which experienced surges of COVID-19 at different times. Figure 4,5,6 show results for California, for CDC, COVID-19 Projections and Covidestim data, respectively. Figures 7,8,9 provide results for New York and Figures 10,11,12 show results for Minnesota.

**Figure 4.**
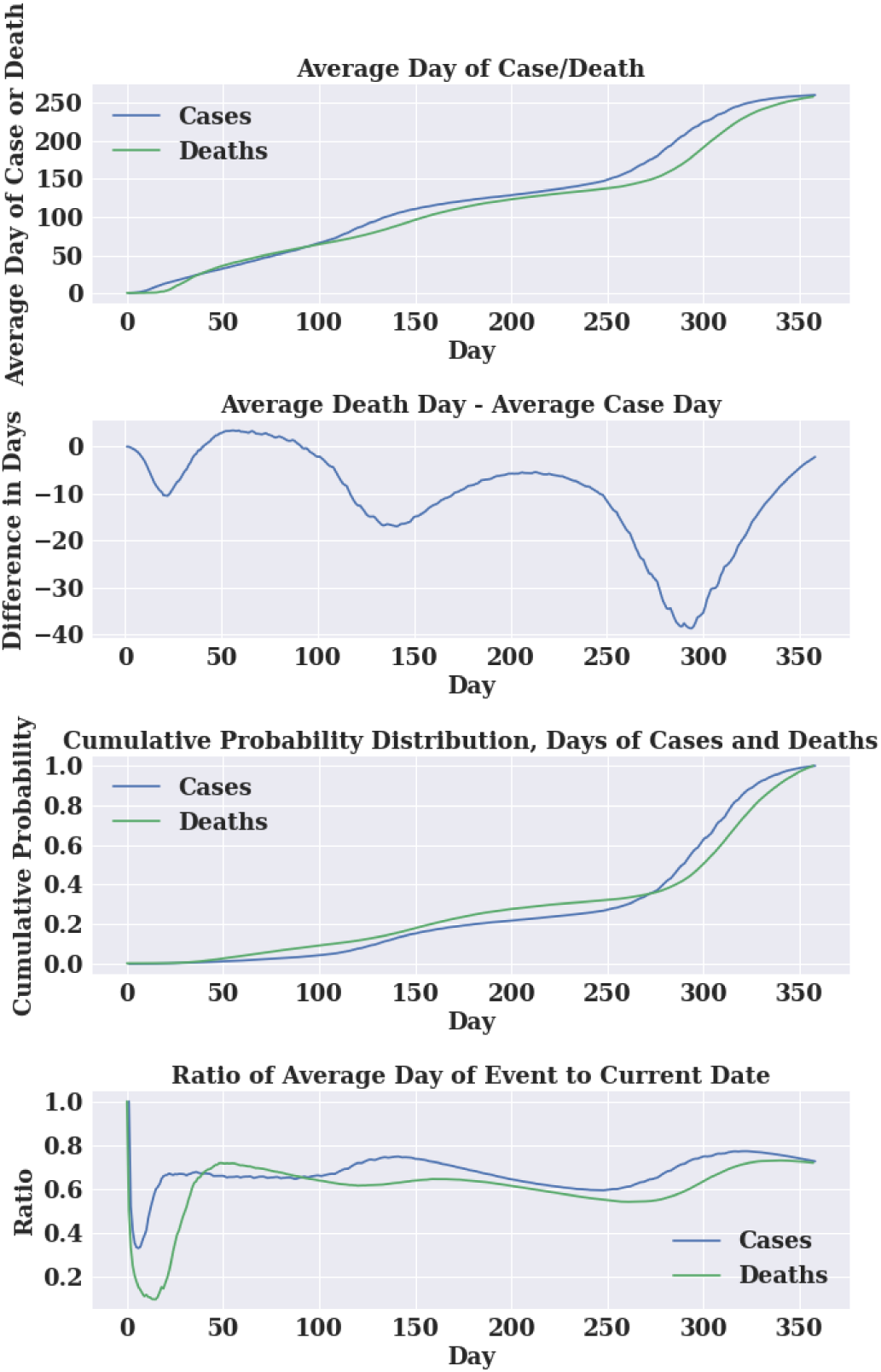
Average Day of Event in California using CDC’s Data

**Figure 5.**
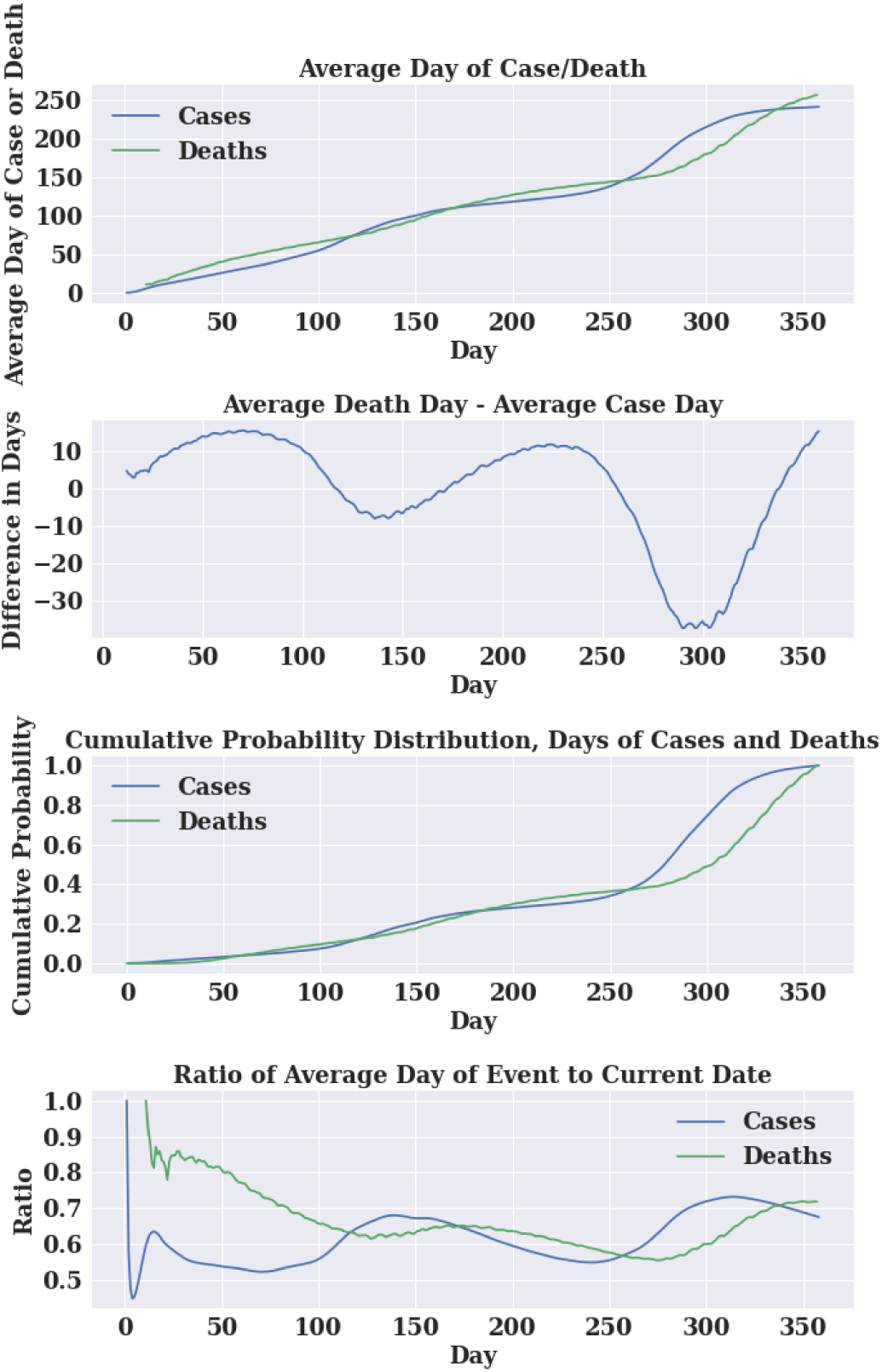
Average Day of Event in California using COVID-19 Projection’s Data

**Figure 6.**
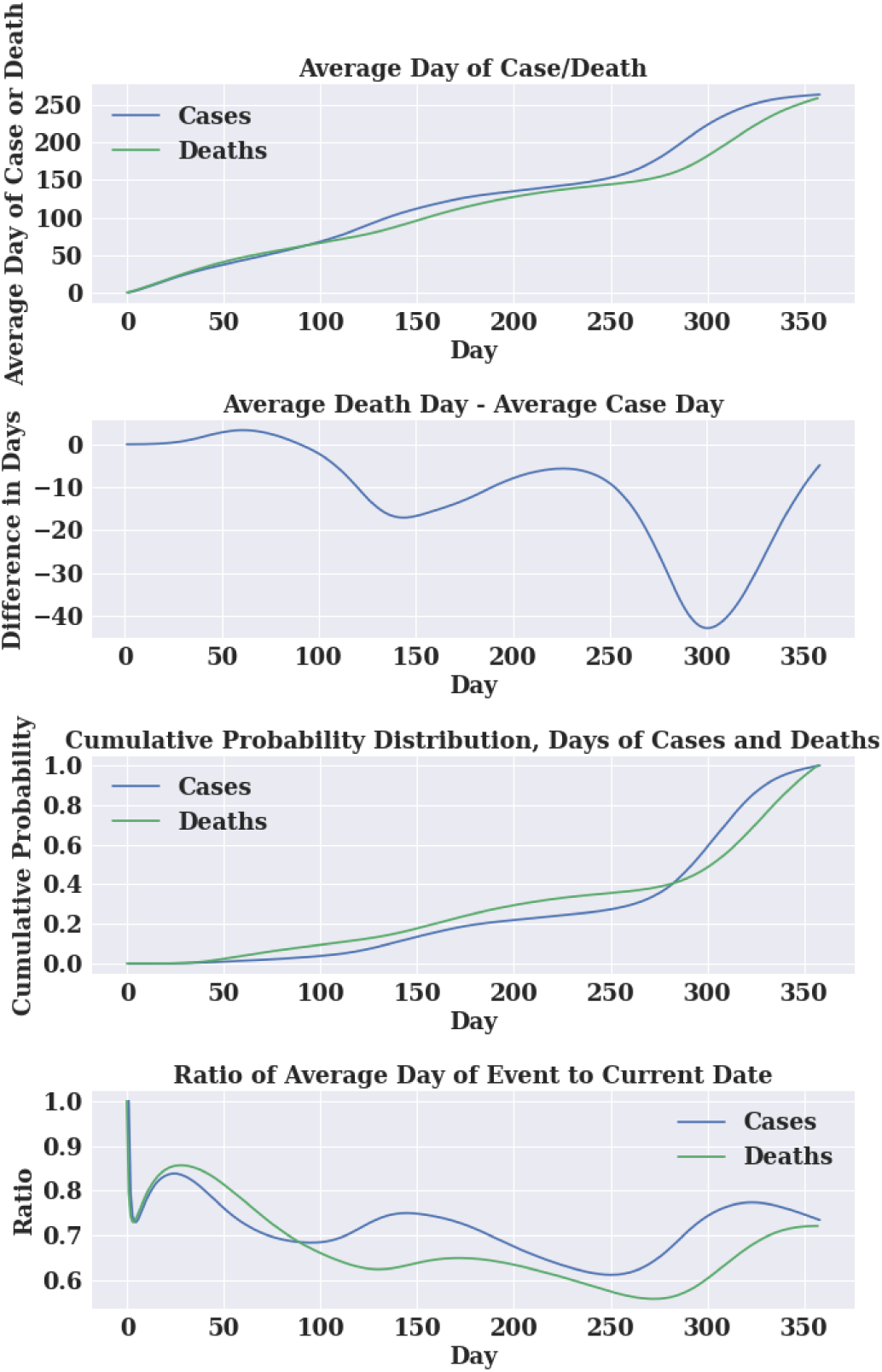
Average Day of Event in California using Covidestim’s Data

**Figure 7.**
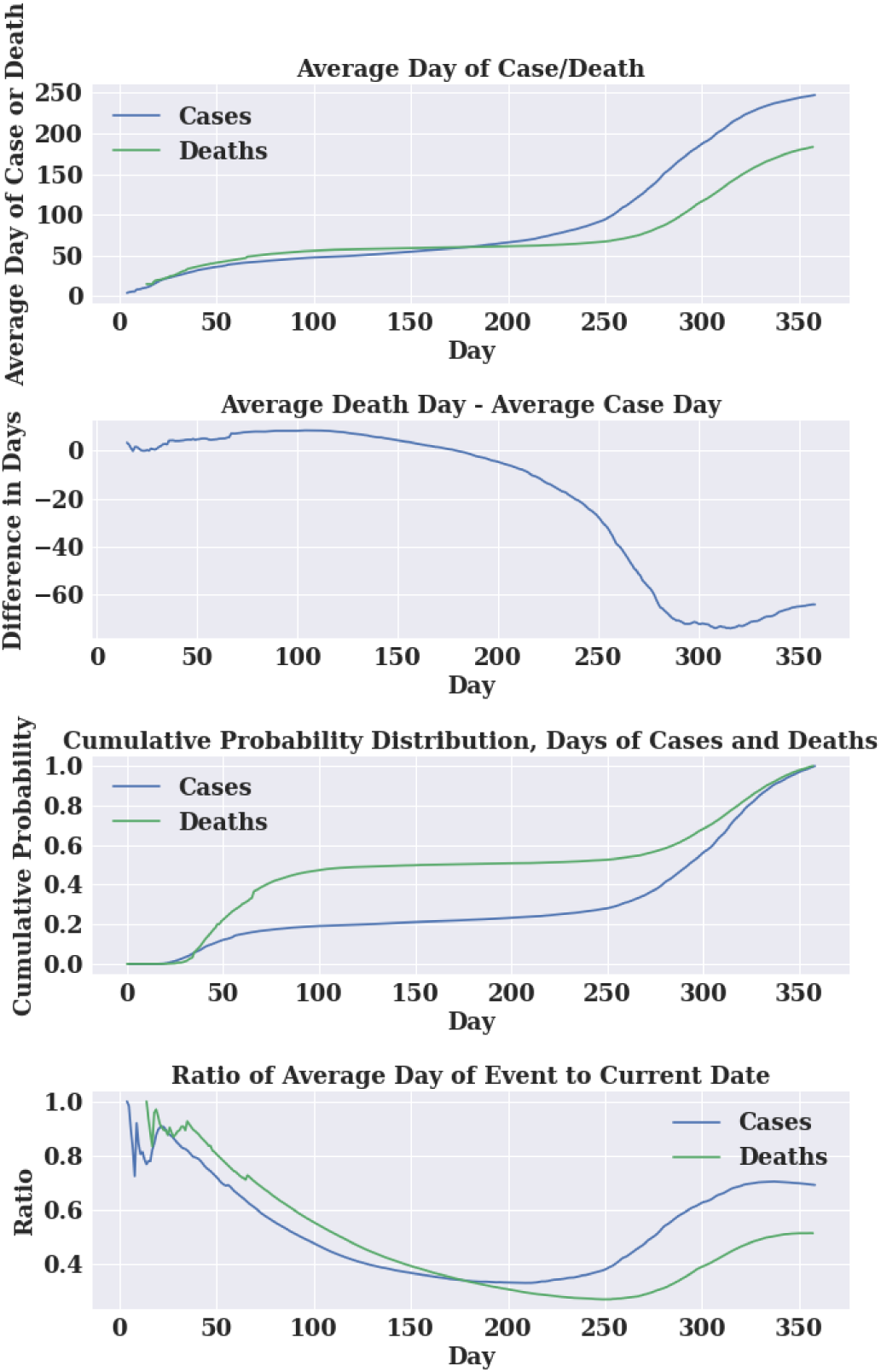
Average Day of Event in New York using CDC’s Data

**Figure 8.**
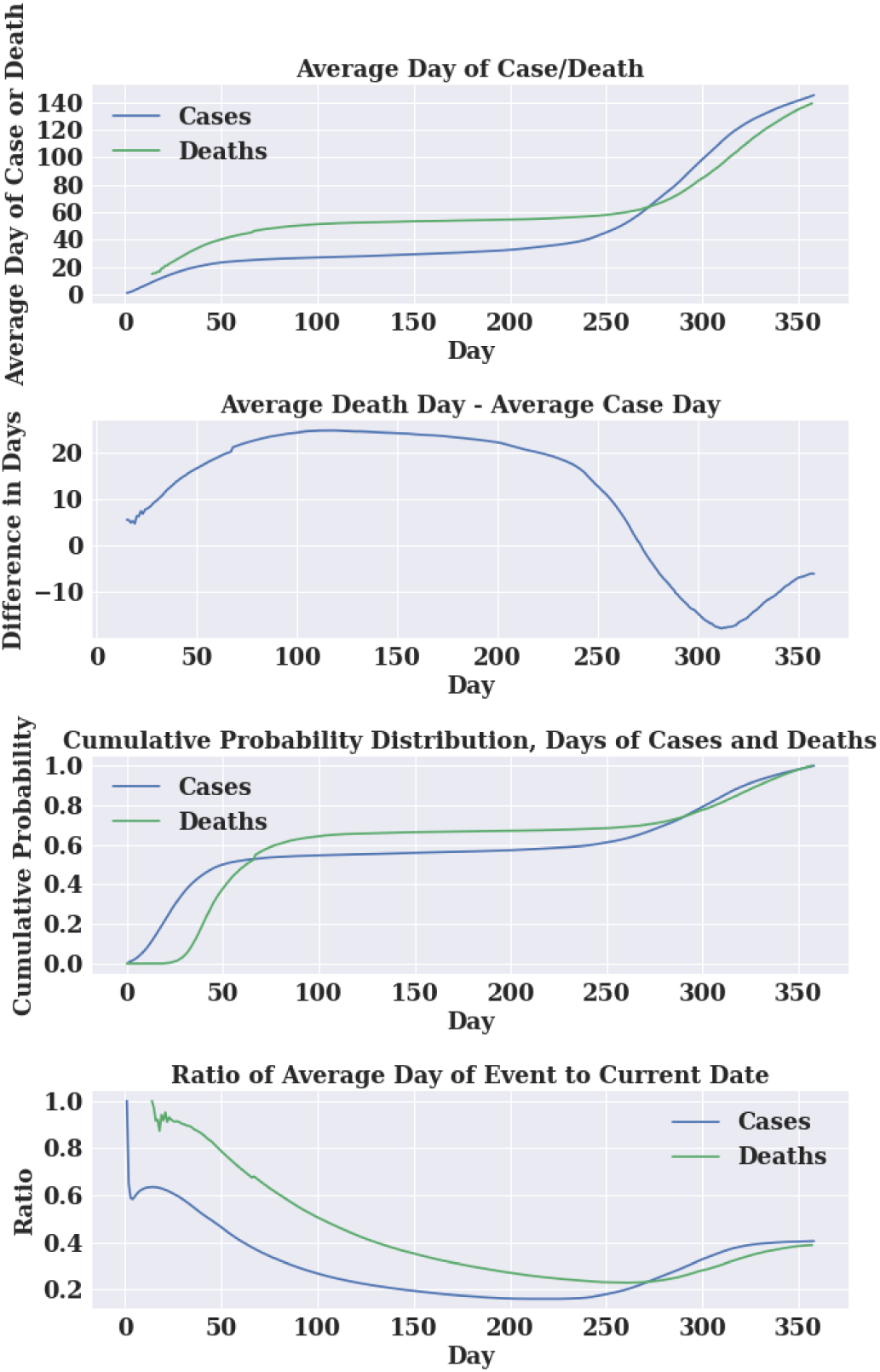
Average Day of Event in New York using COVID-19 Projection’s Data

**Figure 9.**
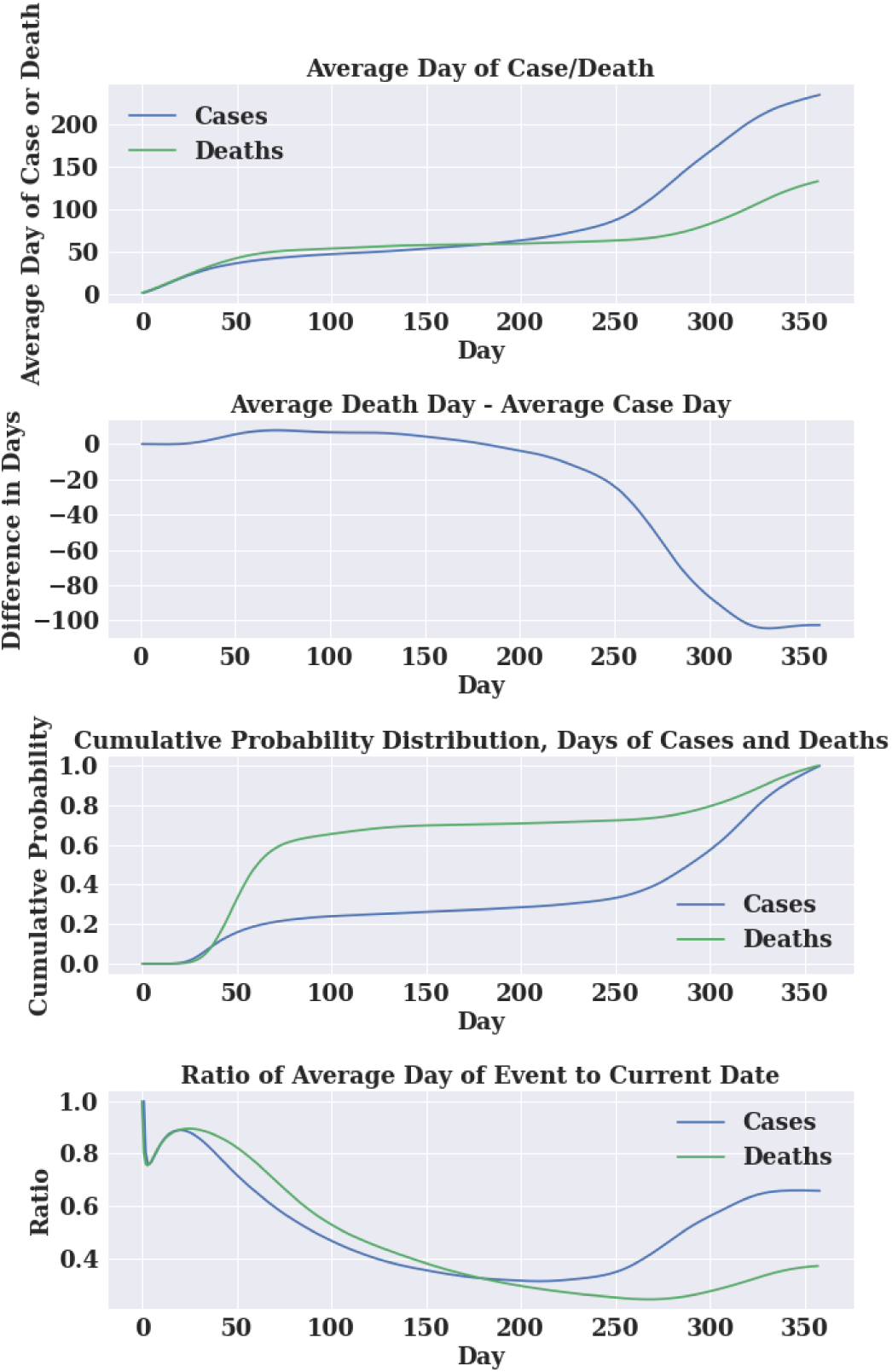
Average Day of Event in New York using Covidestim’s Data

**Figure 10.**
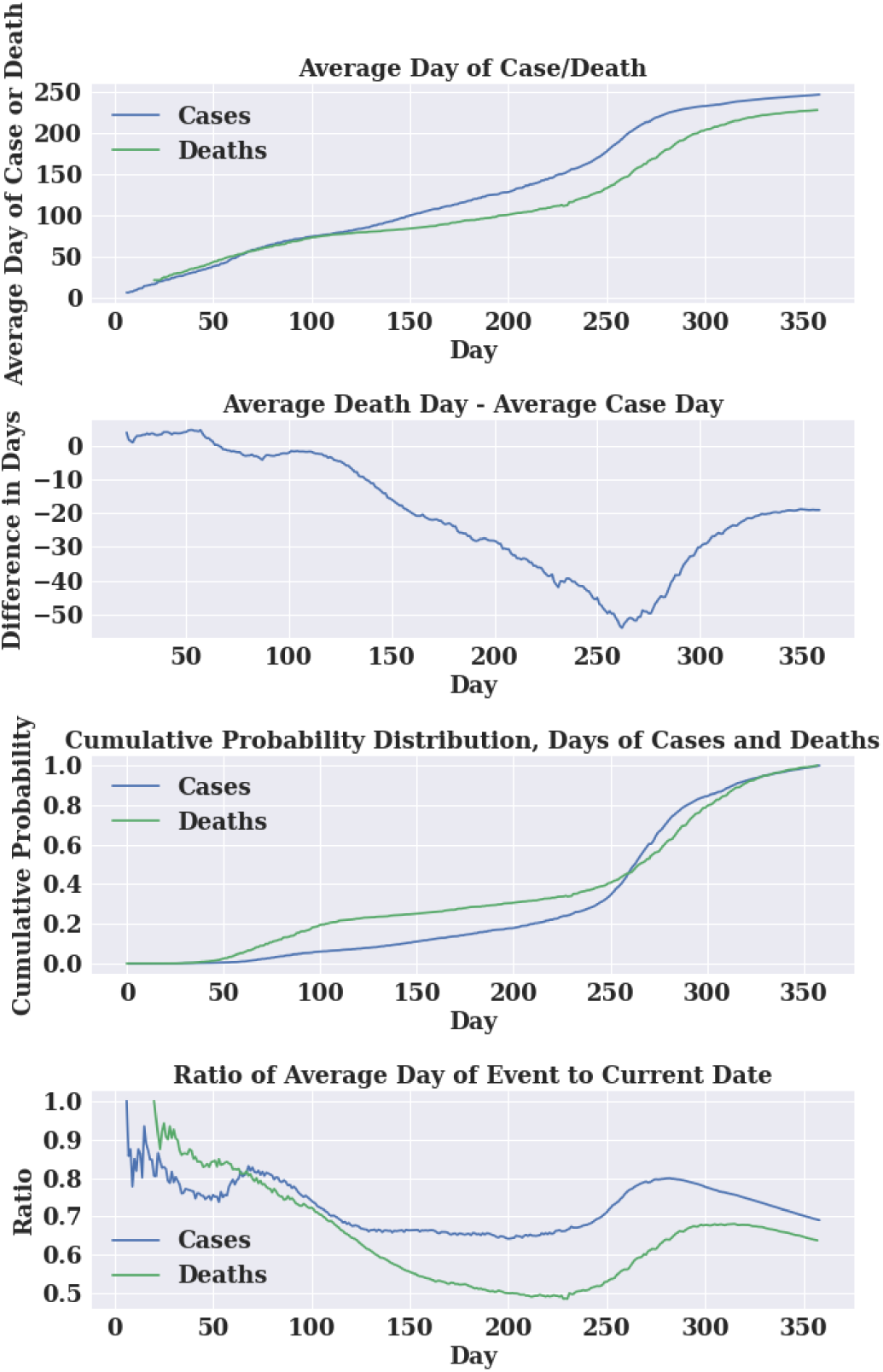
Average Day of Event in Minnesota using CDC’s Data

**Figure 11.**
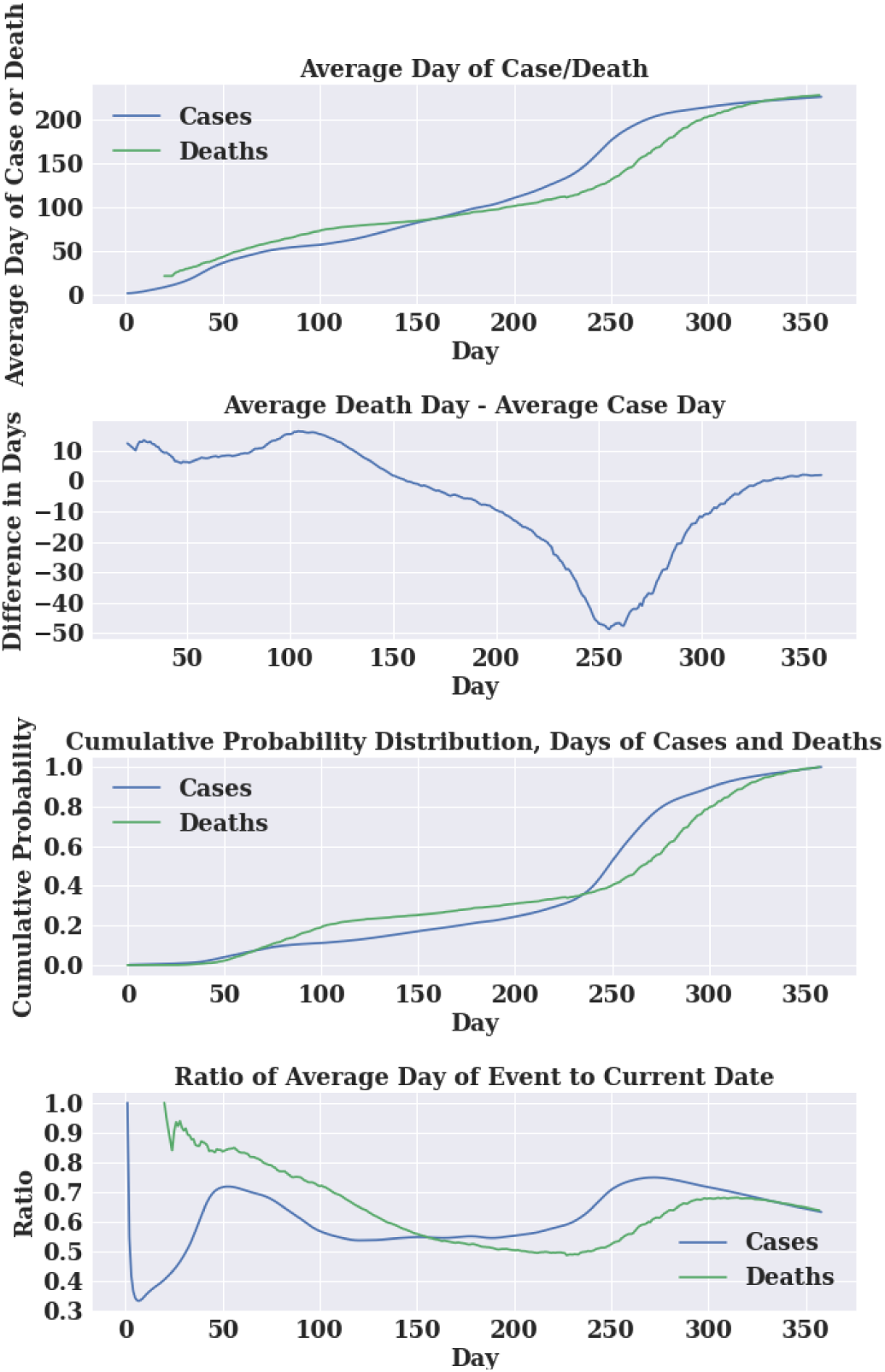
Average Day of Event in Minnesota using COVID-19 Projection’s Data

**Figure 12.**
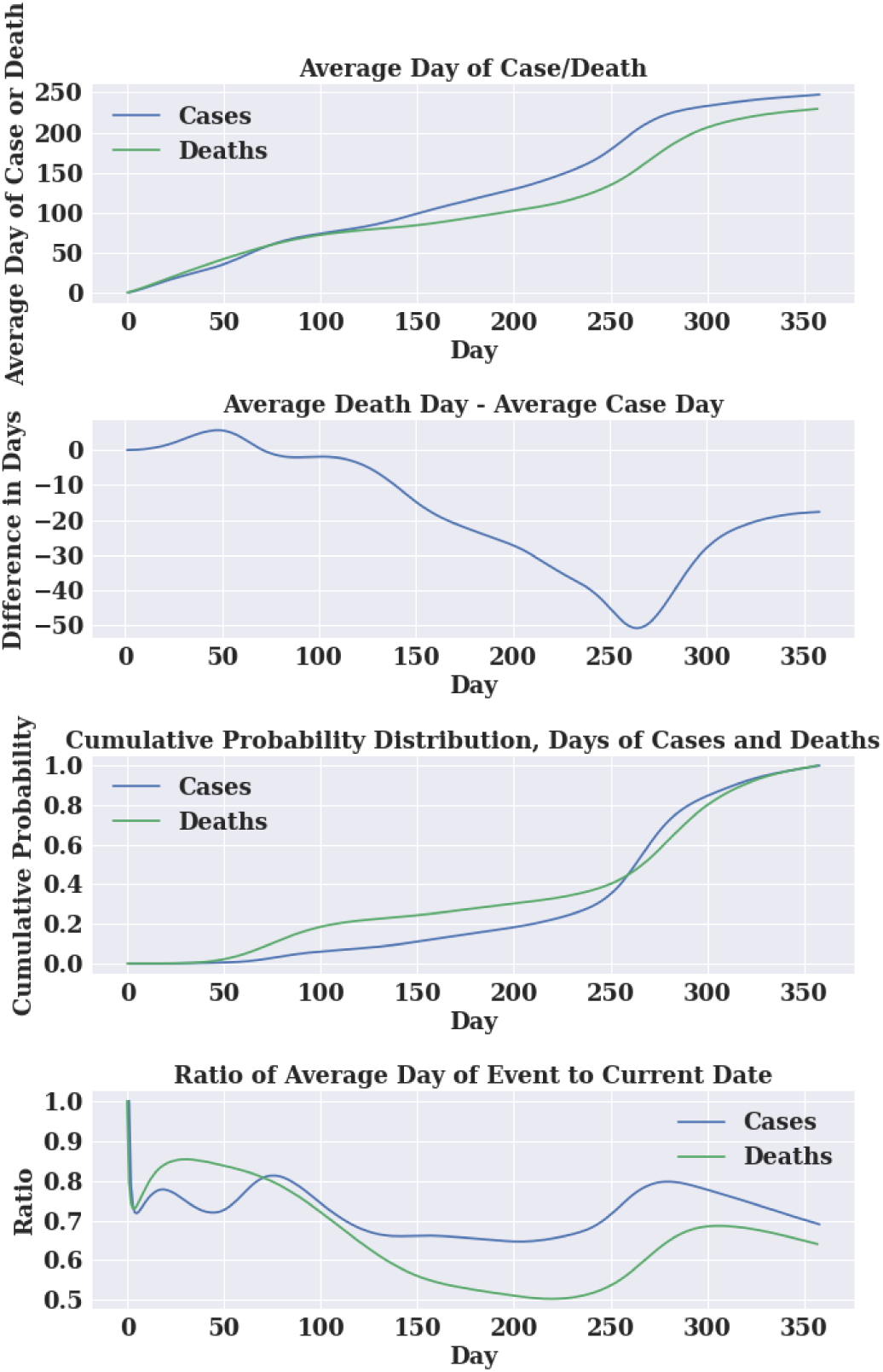
Average Day of Event in Minnesota using Covidestim’s Data

Examining the slopes of *A*_*d*_(*t*) and *A*_*c*_(*t*), geometric growth can be seen in the first 50 days of the pandemic, where these functions increase at an approximate rate of one per day. Between day 50 and 250, *A*_*d*_(*t*) and *A*_*c*_(*t*) increases at slow rates, as reflected in dropping values of *R*_*d*_(*t*) and *R*_*c*_(*t*), indicators that the pandemic waned during that time period. That pattern reversed between day 250 and 300, when *A*_*d*_(*t*) and *A*_*c*_(*t*) increased at super-geometric rates, exceeding one per day, as a second wave of the pandemic began to overwhelm the magnitude of the first wave, at least in California and Minnesota. Super-geometric growth did not occur in death data for New York, owing to the large magnitude of the first wave of COVID-19 experienced in New York.

One would expect that cumulative probability distributions for deaths and cases would follow similar patterns, with deaths lagging cases, unless there have been notable changes in survival rates over time. Yet plots of *L*(*t*) fell well below zero, ranging between -10 and -100 around day 300, before rising at the end of the study period. Other than possible improvement in survival, there are two explanations. First, cases were relatively under-reported at the start of the pandemic, thus creating an upward bias in the dates of reported cases. This is particularly evident in CDC data for New York. Second, in the later wave of the pandemic, cases appeared prior to deaths, thus creating a temporary dip in the *L*(*t*) indicator, as apparent in California and Minnesota.

Comparing the three data sources, measures of deaths are similar, but measures of cases differ significantly. Under COVID-19 projections, cases are distributed earlier in the pandemic, particularly in New York, more closely following the patterns of deaths, which were perhaps a more accurate indicator of actual infections.

### Comparison of Statistics for all 50 States

We now examine comparative statistics for the 50 states. For each state, we calculated the average case day and average death day, as of 2/21/2021, for all three data sources. The distributions of these statistics were then plotted as cumulative distributions in Figures 13 and 14. As shown in Figure 13, average case day, computed from Covidestim, varies from 229 (Louisiana) to 280 (Wyoming), with an average (of the averages) of 253 (Table 1). For contrast, case averages from COVID-19 Projections range from 139 (New Jersey) to 283 (Wyoming) with an average (of the averages) of 227. Thus, the latter estimates show cases occurring much earlier in states that experienced the most severe outbreaks in the March/April timeframe, yet only 26 days earlier on average. The distributions of average death day are similar to COVID-19 Projections’ case day distribution, with averages ranging from about 133 to 283.

**Table 1.** Statistics for Average Case and Death Day by State

|  | Covidestim |  | Covid-19 Projections |  | CDC |  |
| --- | --- | --- | --- | --- | --- | --- |
|  | Average Day of Case in State | Average Day of Death | Average Day of Case in State | Average Day of Death | Average Day of Case in State | Average Day of Death |
| National Average | 253 | 236 | 227 | 235 | 252 | 235 |
| SD Among States | 12 | 34 | 24 | 33 | 12 | 31 |
| Minimum Among States | 230 | 133 | 145 | 139 | 219 | 144 |
| Maximum Among States | 280 | 282 | 264 | 283 | 280 | 283 |

**Figure 13.**
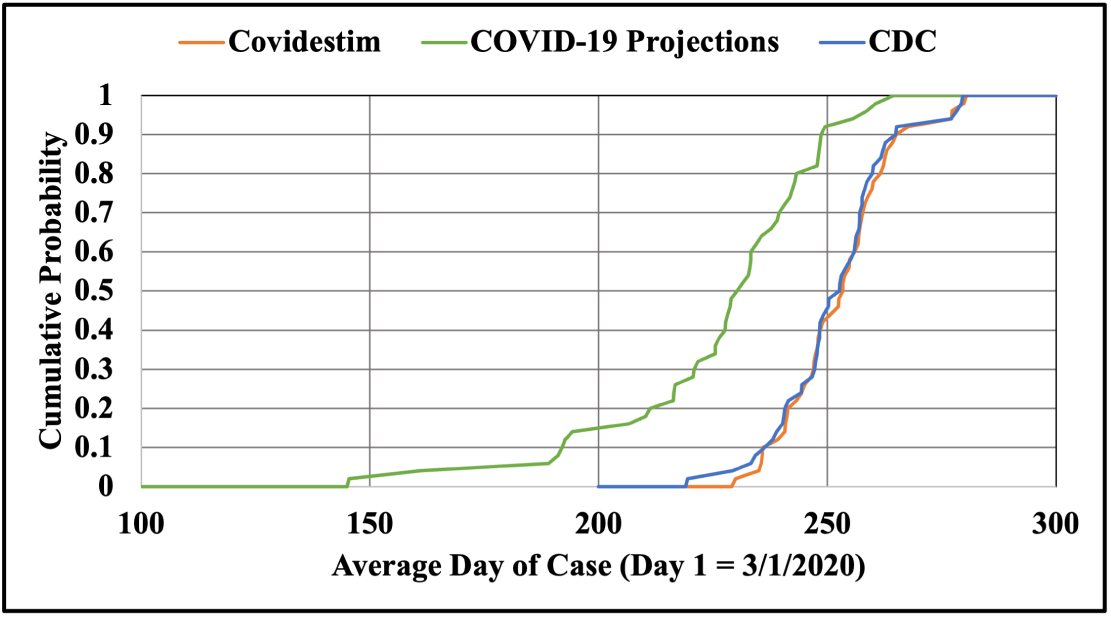
Cumulative Probability Distribution for Average Day of Case Among States

**Figure 14.**
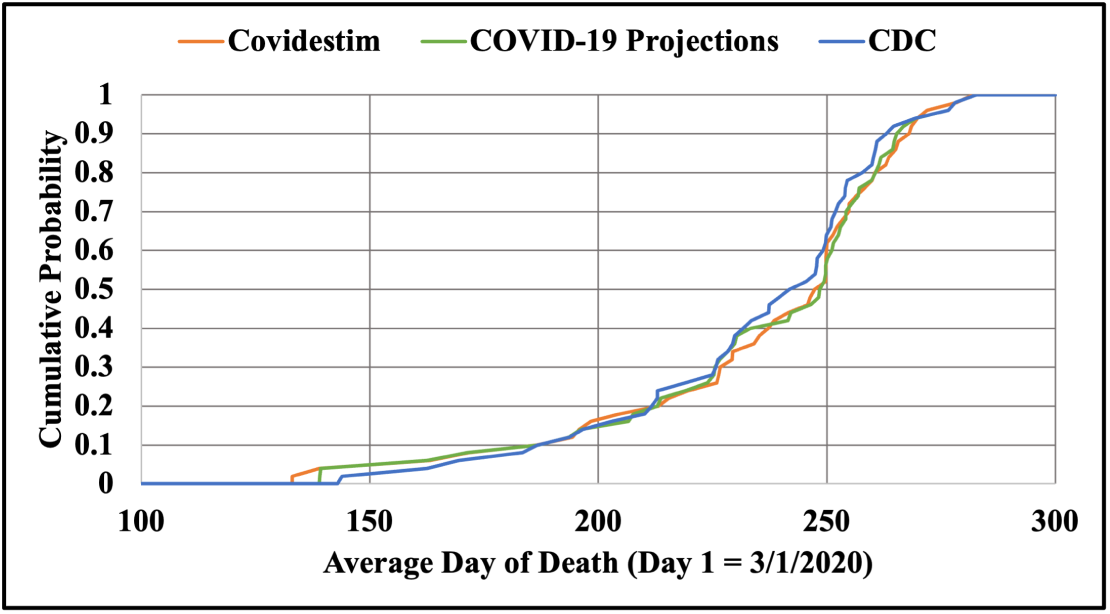
Cumulative Probability Distribution for Average Day of Death Among States

Figure 15 shows how *L*(*t*) varied from month to month. For each month and state, we counted the number of days that *L*(*t*) was positive (i.e., average death day is later than average case day) and then computed an average for this statistic among the states. These results are shown for the three data sources, expressed as the percentage of days within the month. The percentages peaked in April and May, toward the end of the first wave of the pandemic. They dropped to a minimum toward the peak of the December wave, as cases appeared prior to associated deaths. However, even as the pandemic waned in February, the average case day tended to be later than the average death day (though less so with COVID-19 Projections).

**Figure 15.**
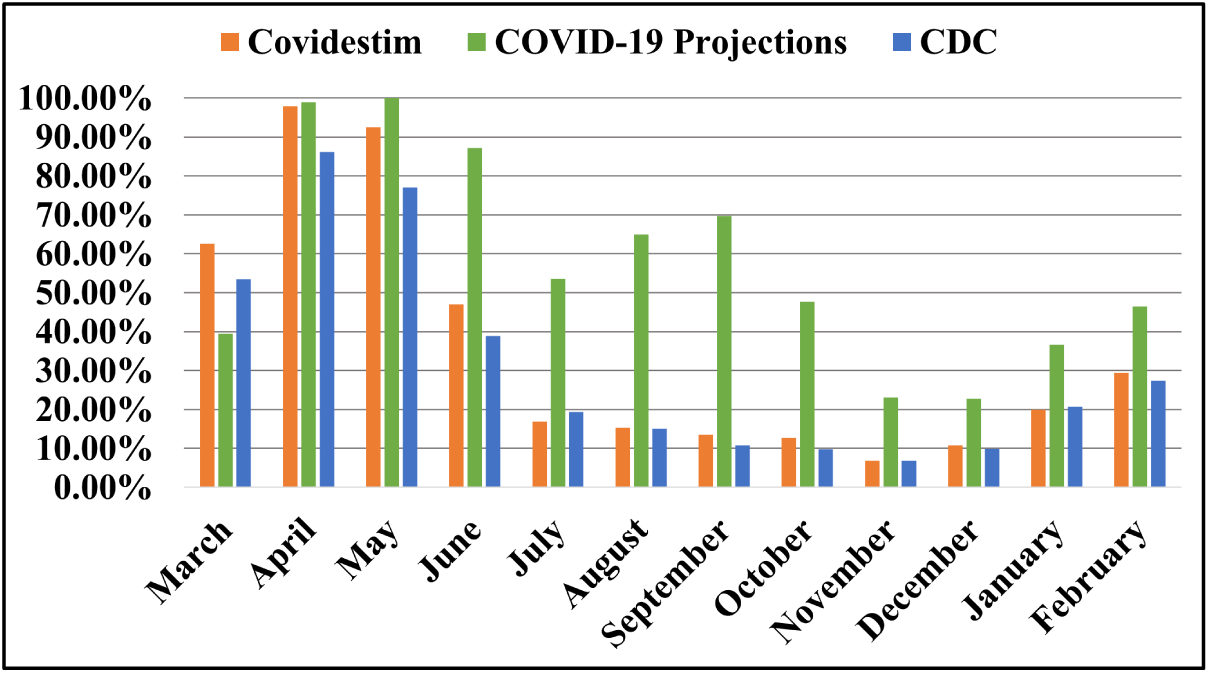
Average % of Days that Average Death Day is Greater than Average Case Day (March 2020 to February 21, 2021)

Figures 16,17,18 provide another graphical comparison among states, which are color-coded as to the average case days for CDC and COVID-19 Projections data, and average death day for CDC data. Notably, CDC case data suggest a much narrower distribution for the time distribution of the pandemic than either CDC death data or COVID-19 Projections case data, for which the pandemic is shown to occur much earlier in New York and surrounding states, as well as in Louisiana.

**Figure 16.**
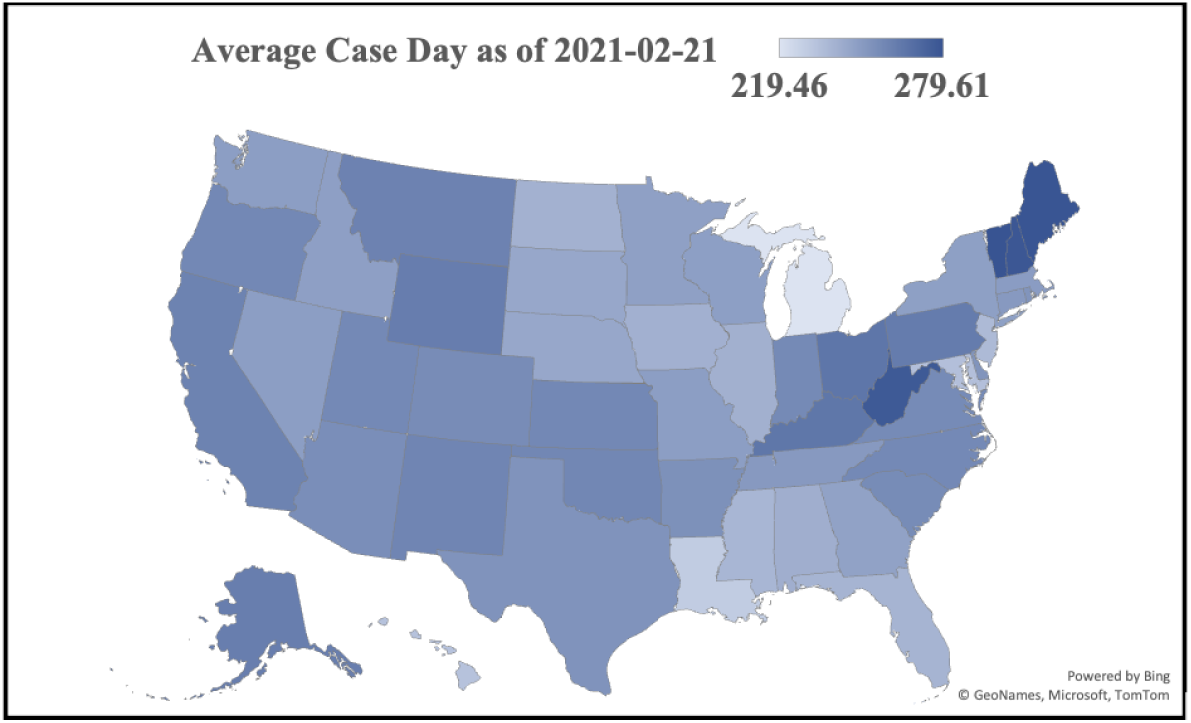
Average Case Day on 2021-02-21 (CDC)

**Figure 17.**
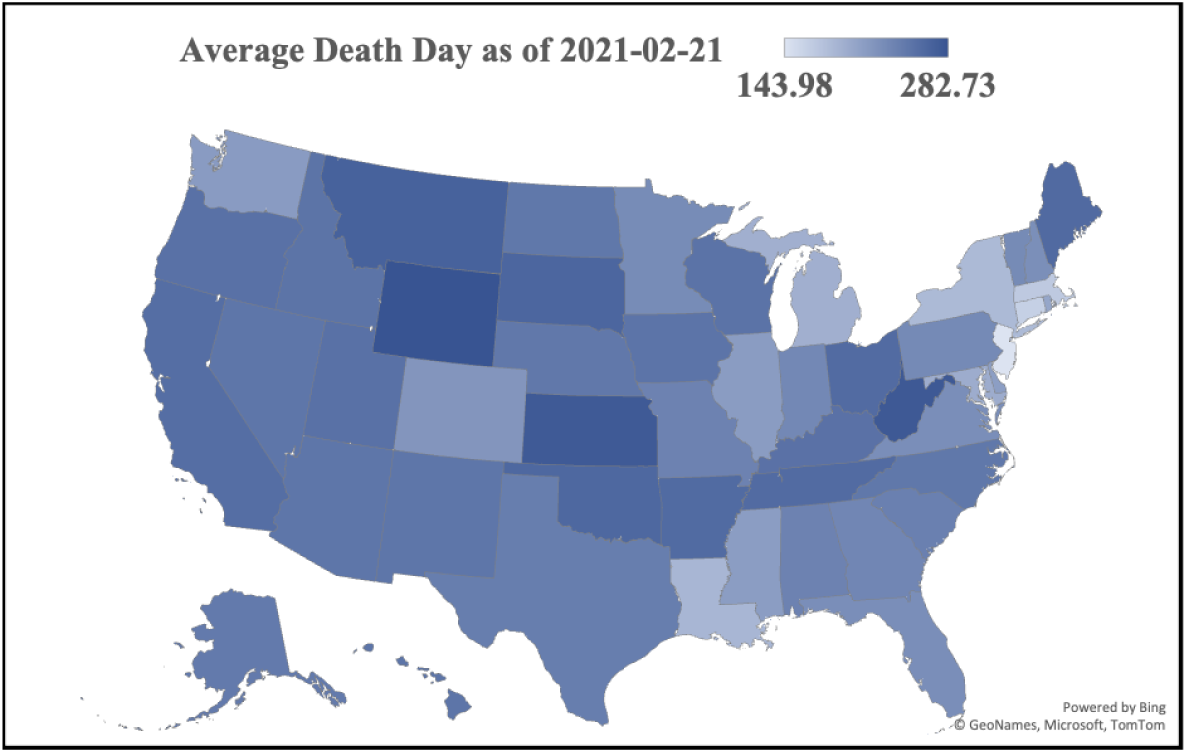
Average Death Day on 2021-02-21 (CDC)

**Figure 18.**
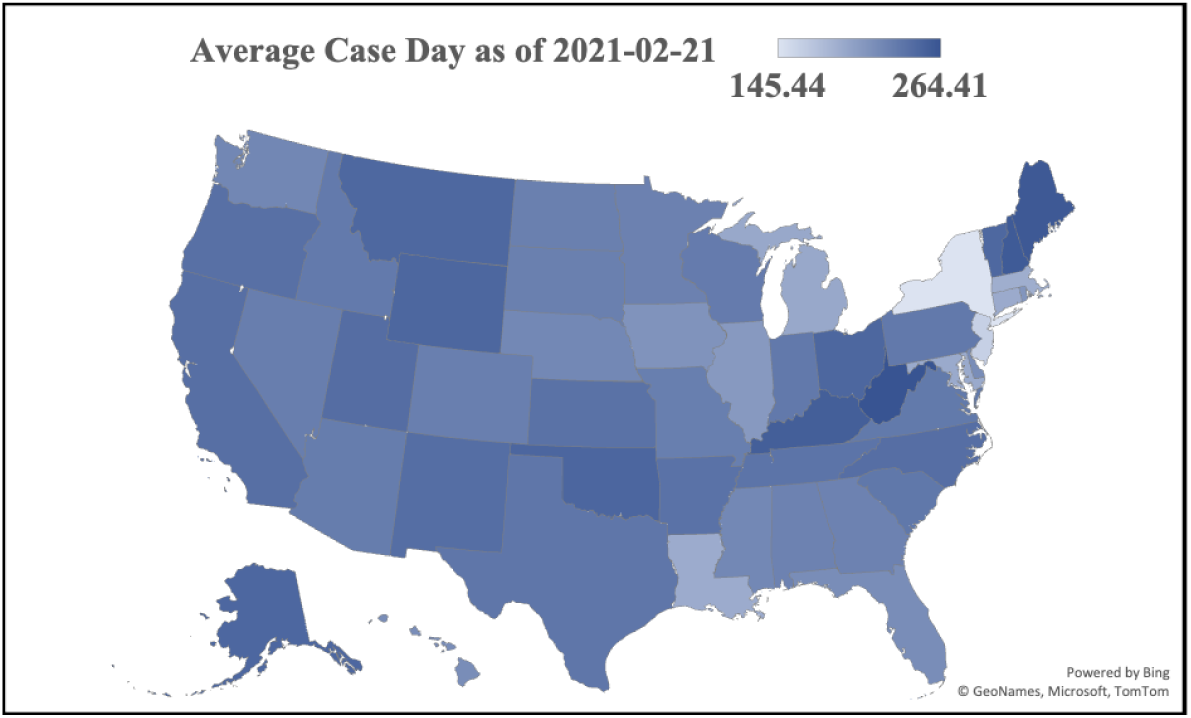
Average Case Day on 2021-02-21 (COVID-19 Projections)

Figure 19 provides a final comparison, plotting the average day of case minus the average day of death (which we call the difference in days) for CDC data. Each dot represents a state, which are color coded by region (according to US Census designation) and plotted according to state population (2020 census). Visually, the difference in days appears largest in northeastern states. By linear regression, the difference in days was only found to be significantly different in the northeast region than other regions in the country (55 days on average, with t statistic of 6.5). State population was not a significant determinant of the difference in days (t=1.0). These results appear to reflect the early experience of COVID in northeastern United States, when reported deaths were proportionately much higher relative to reported cases compared to later in the pandemic.

**Figure 19.**
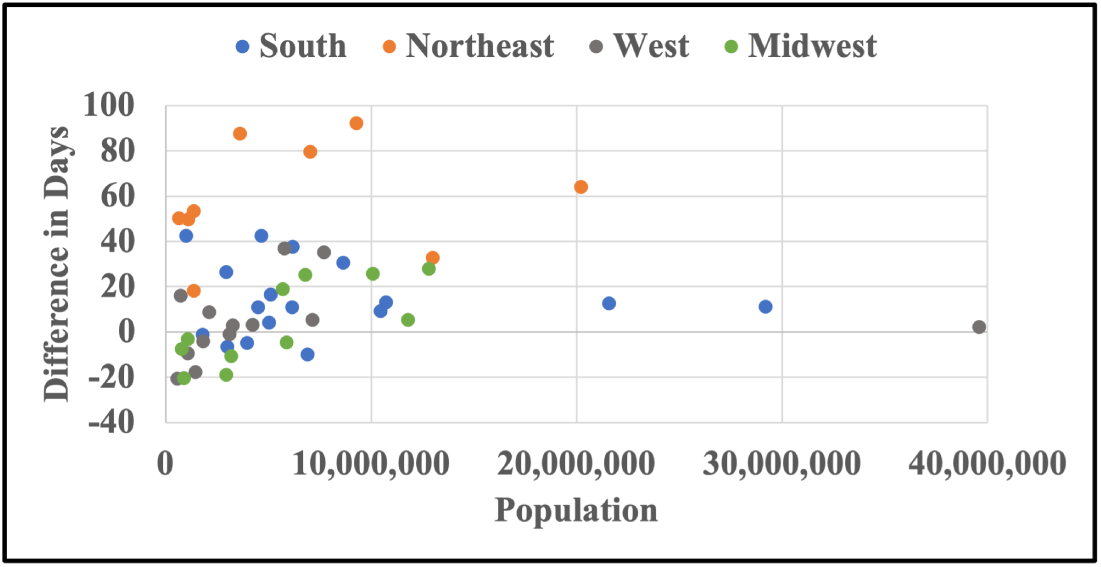
Average Case Day on 2021-02-21 (COVID-19 Projections)

## Discussion and Conclusions

We have developed and applied a methodology to illustrate how the time distribution of disease events (cases and deaths) varies over time by location. We see how geometric growth of daily cases and deaths result, in the limit, to a constant separation between the present day and the average event day, and we see how this phenomenon appeared in the early days of the pandemic, particularly in New York. When the partial average grows at the rate of one per day, the pandemic is accelerating. We also found that later in the pandemic, during a second or third wave, the partial average grew at rates exceeding one per day. Thus, the separation between the current day and the average day became smaller, signs that the pandemic worsened. We also observed that the time distribution varied by several months among the 50 states, as the pandemic accelerated and decelerated at different times. Last, we observed that the average day of deaths preceded the average day of cases much of the time, particularly during the December 2020 to January 2021 period.

The key contribution of this paper has been to provide a normalized representation through which the time distribution of cases and deaths can be compared, by location and data source. These representations supplement other conventional measures in a way that provides new insights into both the accuracy of underlying data and the actual state of pandemic by locality. The analyses might then be applied toward implementing interventions that reflect the trajectory of the disease, and to retrospectively analyze the effectiveness of interventions that may correlate with historical data.

## Data Availability

All the data is available on the website and could be used under permission.

https://covid19datasource.usc.edu/

## Acknowledgments

Research was supported by the University of Southern California through the Zumberge Innovation Fund and the Center for Undergraduate Research in Viterbi Engineering (CURVE).

